# Real-world perioperative FLOT delivery in gastric and gastro-oesophageal junction adenocarcinoma: a single-centre service evaluation in a District General Hospital in the UK

**DOI:** 10.64898/2026.10.19.26365836

**Authors:** Riya Sabharwal, Divyalakshmi Bhaskaran

**Affiliations:** KING’S COLLEGE HOSPITAL NHS FOUNDATION TRUST; St James’s University Hospitals, Leeds Teaching Hospitals, NHS Trust, Beckett Street, LS9 7TF

**Keywords:** FLOT chemotherapy, gastric cancer, gastro-oesophageal junction cancer, relative dose intensity, chemotherapy toxicity

## Abstract

**Background:** Although perioperative fluorouracil, leucovorin, oxaliplatin and docetaxel (FLOT) is the standard neoadjuvant regimen for resectable gastric and gastro-oesophageal junction adenocarcinoma, evidence describing treatment delivery in routine clinical practice remains limited, particularly regarding relative dose intensity (RDI) and treatment completion.

**Objectives:** To evaluate real-world delivery of preoperative FLOT chemotherapy, including treatment completion, progression to surgery, RDI, toxicity and exploratory changes in estimated lean body mass (LBM).

**Methods:** A retrospective single-centre service evaluation included consecutive patients receiving neoadjuvant FLOT during 2024. Outcomes included treatment completion, surgery, RDI, toxicity, pathological response and estimated LBM. Descriptive statistics were used.

**Results:** Twelve patients were included (median age 68.6 years; 91.7% male). Eleven (91.6%) completed all four preoperative cycles and proceeded to surgery. Mean RDI was 76.7%. Pre-emptive dose capping was the predominant dose modification strategy, with only one reactive dose reduction. Two grade 3 adverse events occurred. Major pathological response (Mandard grade 1–2) was observed in 2/12 patients (16.7%). Mean estimated LBM declined from 54.3 to 53.4 kg. Greater LBM loss appeared to coincide with poorer clinical outcomes.

**Conclusions:** High treatment completion and progression to surgery were achieved despite reduced RDI. These findings support prospective evaluation of strategies to optimise chemotherapy delivery while maintaining tolerability in routine clinical practice.

**Key Messages:** *What is already known on this topic?:* Perioperative FLOT is standard treatment for resectable gastric and gastro-oesophageal junction adenocarcinoma, but toxicity frequently necessitates dose modification. Real-world data on relative dose intensity and treatment completion remain limited.

*What this study adds?:* In this single-centre cohort, 91.6% of patients completed all four planned preoperative FLOT cycles and proceeded to surgery despite a mean relative dose intensity of 76.7%. Pre-emptive dose capping was the predominant dose-modification strategy.

*How this study might affect research, practice or policy?:* Routine monitoring of relative dose intensity alongside treatment completion may provide a useful measure of chemotherapy delivery. Larger studies should evaluate whether individualised dose optimisation can maintain treatment continuity while balancing toxicity and dose intensity.

## Introduction

Perioperative FLOT (fluorouracil, leucovorin, oxaliplatin, and docetaxel) is the established standard of care for locally advanced resectable gastric and gastroesophageal junction (GOJ) adenocarcinoma treated with curative intent.[1] The FLOT4-AIO trial demonstrated improved overall survival compared with the epirubicin-based regimen ECF/ECX (median 50 vs 35 months; HR 0.77, p=0.012), alongside higher rates of pathological complete response and R0 resection.[1] These results have been incorporated into international guidelines and have shaped contemporary curative-intent practice worldwide.[2]

Although FLOT is established as standard treatment, evidence describing its delivery in routine clinical practice remains limited. [3] Real-world populations frequently differ from trial cohorts, comprising older patients with greater comorbidities reduced physiological reserve, with variable pharmacodynamics and pharmacokinetics. Understanding FLOT delivery in everyday practice, and how this relates to treatment completion, toxicity, and pathological outcomes, impacts optimisation of care and identifying opportunities for sustaining durable responses to systemic anticancer treatments (SACT) while reducing toxicity burden.

The efficacy of FLOT is accompanied by significant toxicity. In the FLOT4-AIO trial, grade 3–4 neutropenia occurred in approximately 50% of patients, grade 3–4 infections in 18%, and grade 3–4 diarrhoea in 10%.[1] Notably, only 46% of trial participants completed all eight planned cycles, though 94% proceeded to surgery.[1] Relative dose intensity (RDI) provides a well-established metric for evaluating SACT delivery. Defined as the ratio of delivered to planned dose intensity per unit time, RDI has been widely adopted as a quality measure in curative-intent chemotherapy, with thresholds of ≥85% generally recommended for optimal outcomes. [4,5,6,7] In routine practice, however, the balance between maintaining dose intensity and preserving tolerability requires careful clinical judgement, particularly in patients whose reserve may be more limited than that of trial participants.

Optimising chemotherapy delivery requires careful balancing of dose intensity, toxicity and progression to surgery. Dose modification may be required to maintain treatment tolerability while preserving curative intent, although evidence describing how these decisions are implemented in routine practice remains limited. Proactive dose modification, anticipating toxicity rather than responding to it, represents one strategy for preserving treatment continuity while minimising treatment-related morbidity. [8] Standardised documentation of toxicity and dose adjustments, facilitated through electronic prescribing systems overseen by pharmacy teams, demonstrated improved consistency and transparency across clinical teams.[9]

Beyond dose intensity, patient physiological reserve influences treatment tolerance. Estimated lean body mass (LBM), calculable from routine anthropometric data using the Janmahasatian formula, may provide an accessible measure of metabolic reserve.[15] Unlike body surface area (BSA), which increases approximately linearly with total body weight, Janmahasatian-derived LBM increases nonlinearly and plateaus at higher body mass indices (BMI), reflecting that excess body weight at higher BMI is predominantly adipose rather than metabolically active tissue. [10,11] Consequently, BSA-based dosing may overestimate the pharmacologically relevant distribution compartment in overweight or obese patients, potentially increasing the risk of toxicity for agents that distribute primarily within lean tissues and intravascular space.[12]

Studies in gastrointestinal malignancies have associated low LBM with increased chemotherapy toxicity, including dose-limiting toxicity during oxaliplatin-based regimens.[12,13,14] CT-derived skeletal muscle analysis remains the reference standard for assessing sarcopenia, although the potential utility of pragmatic anthropometric estimates such as the Janmahasatian formula has not been explored in routine perioperative gastric cancer practice[15]

This study evaluated treatment completion, RDI, toxicity and pathological response during routine perioperative FLOT delivery within an NHS district general hospital, comparing findings with published trial data.

## Aims

### Primary

To evaluate treatment delivery, relative dose intensity and progression to surgery among patients receiving perioperative FLOT.

### Secondary

- dose modifications
- toxicity
- pathological response
- exploratory LBM observations

## Methods

### Design

This retrospective single-centre service evaluation included consecutive patients receiving perioperative FLOT during 2024 (one year data). The evaluation was registered with the hospital clinical governance department (CSCS/ONC/142). Reporting followed the Strengthening the Reporting of Observational Studies in Epidemiology (STROBE) statement and the ESMO Guidance for Reporting Oncology Real-World Evidence (GROW). The ESMO-GROW flow diagram is provided as Supplementary Appendix 1. [18] Descriptive analyses only were performed.

### Population

Thirteen patients treated between January and December 2024 with biopsy-proven gastric or GOJ adenocarcinoma scheduled for perioperative neoadjuvant SACT (FLOT) were identified. One patient who discontinued treatment after a single cycle was excluded. The final analysis therefore included 12 patients. No further exclusion criteria were applied; all patients commencing preoperative FLOT during the study period were captured.

### Data Collection and Integrity

Demographic, treatment, toxicity and pathological data were extracted retrospectively from the electronic patient record and chemotherapy prescribing system into an anonymised dataset for descriptive analysis using Jamovi (version 2.6.44).

#### Relative Dose Intensity

RDI was calculated for each drug within the FLOT regimen. For each agent, the delivered dose intensity (total dose delivered divided by actual treatment duration) was expressed as a proportion of the planned dose intensity (standard dose per cycle divided by the planned cycle interval of 14 days). The overall RDI per patient was taken as the mean of the individual drug RDIs across all FLOT components, accounting for both dose reductions and treatment delays within a single metric.[16]

#### Estimated Lean Body Mass

Estimated lean body mass was calculated using the Janmahasatian equation from routinely recorded height and weight. Changes between baseline and completion of neoadjuvant chemotherapy were analysed descriptively. These analyses were exploratory and not intended as surrogate measures of CT-derived body composition.[17]

#### Toxicity Capture

Toxicity data were extracted from medical, nursing, and chemotherapy administration records. In the absence of prospective Common Terminology Criteria for Adverse Events (CTCAE v6) grading in routine documentation, toxicity grades were assigned retrospectively by the lead auditor; this approach carries an inherent risk of under-reporting, particularly for lower-grade toxicities, and should be considered when interpreting the toxicity findings.

### Analysis

Continuous variables are presented as medians (range) or means (SD), and categorical variables as frequencies (%). No inferential analyses were undertaken.

## Results

### Cohort Characteristics

Twelve patients were included (11 male, 1 female), with a median age of 68.6 years. All had biopsy-proven gastric or GOJ adenocarcinoma and were scheduled for perioperative FLOT with curative intent.

### Pathway Timing

The median interval from diagnosis to commencement of SACT was 41 days, within the NHS 62-day cancer treatment target.

### Cycle Completion and Progression to Surgery

Eleven patients (91.6%) completed all four planned preoperative FLOT cycles; one patient discontinued after cycle 3 due to grade 3 sepsis. Eleven patients (91.6%) proceeded to surgery, compared with 94% in the FLOT4-AIO trial (figure 1).[1]

**Figure 1.**
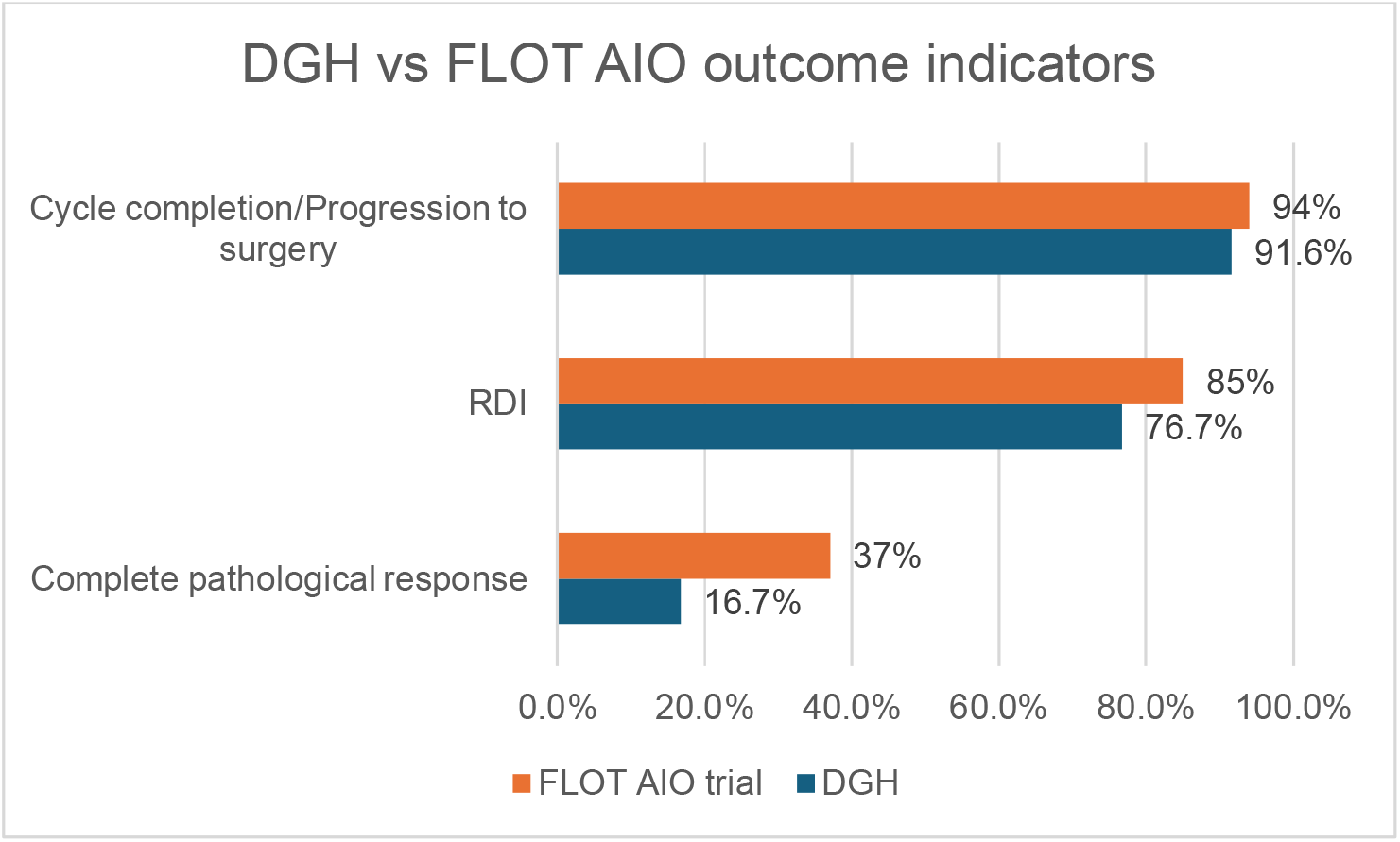
Comparison of treatment delivery and pathological outcomes between the study cohort and the FLOT AIO trial. Abbreviations: DGH, district general hospital; FLOT4-AIO, fluorouracil, leucovorin, oxaliplatin and docetaxel versus epirubicin-based chemotherapy trial; RDI, relative dose intensity; TRG, tumour regression grade.

### Dose Intensity

The mean cohort RDI was 76.7 (14.7)%, below the conventionally recommended threshold of ≥85%(figure 1).[4,5,6,7] The predominant pattern was pre-emptive dose capping rather than reactive dose reduction: 10 patients commenced SACT at reduced dose levels based on clinical assessment of anticipated tolerance, with one patient beginning at 60% of protocol dose. Only one patient required a reactive dose reduction during treatment, following grade 3 fatigue. This pattern is consistent with a proactive approach to dose modification aimed at preserving treatment continuity and fitness for surgery.[8]

### Biometrics

Mean BMI decreased modestly from 27.2 at baseline to 26.6 while mean estimated LBM decreased from 54.3 kg at baseline to 53.4 kg post-treatment, representing a mean reduction of 1.7%. The percentage changes in LBM (LBM_delta_percent) are shown in figure 2.

**Figure 2.**
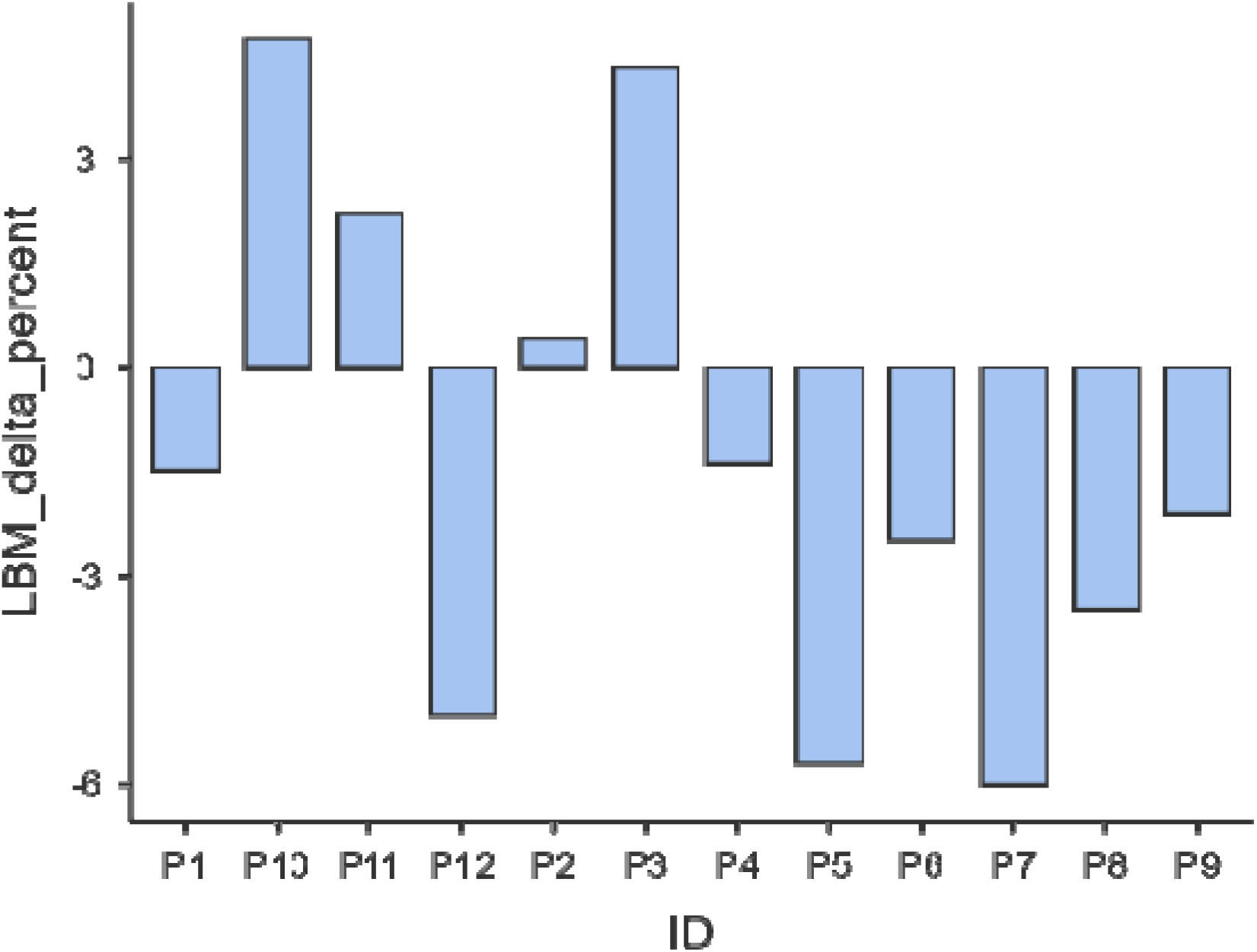
Individual percentage change in estimated lean body mass following preoperative FLOT chemotherapy. Abbreviations: LBM, lean body mass.

### Toxicity

Grade 3 adverse events were documented in two patients: one episode of grade 3 sepsis leading to early SACT discontinuation, and one episode of grade 3 fatigue prompting reactive dose reduction.

### Pathological Response

Major pathological response, defined as Mandard tumour regression grade 1–2, was observed in two patients (16.7%). This is approximately half the rate reported in the FLOT4-AIO trial (37%)(figure 1).[19]

### Discussion

Preoperative FLOT delivery at this DGH achieved high rates of cycle completion (91.6%) and progression to surgery (91.6%), closely matching the FLOT4-AIO benchmark of 94%, despite a mean delivered RDI of 76.7%, below the conventionally recommended threshold of ≥85%.[1,5,6,7] Given the single-centre design and small sample size, this observation is descriptive and no inference regarding pathway performance can be drawn from this alone. Nevertheless, the high proportion of patients completing neoadjuvant SACT and proceeding to curative-intent surgery raises important questions about how dose intensity, treatment completion, and clinical outcomes interact in routine practice.

The coexistence of high completion rates alongside reduced dose intensity reflects a deliberate clinical strategy. In this cohort, pre-emptive dose capping was the predominant approach; several patients commenced SACT at reduced doses based on individualised assessment of anticipated tolerance, and one patient began at 60% of protocol dose. Only one patient required reactive dose reduction due to grade 3 fatigue, and one patient discontinued early following grade 3 sepsis. This proactive approach appeared to facilitate completion of the planned neoadjuvant course and preserve fitness for surgery in this small patient group, consistent with emerging evidence that pre-emptive dose modification strategies may maintain treatment continuity while minimising severe toxicity.[8]

The finding that RDI fell below 85% in this cohort raises a clinically important question: whether higher dose delivery could be safely facilitated in appropriate patients through more proactive and structured supportive care strategies, rather than through pre-emptive dose reduction alone. Several evidence-based approaches warrant consideration in this regard.

Comparison with trial outcomes warrants careful interpretation. The tumour regression grade (Mandard TRG 1–2) of 16.6% was approximately half the 37% reported in the FLOT4-AIO trial.[19] This difference should be interpreted cautiously because of the small sample size, differences between trial participants and routine NHS populations, and potential variability in pathological assessment. Whether reduced RDI contributed to lower pathological response cannot be determined from this descriptive study and warrants evaluation in larger real-world cohorts. Exploratory analysis suggested that greater reductions in estimated LBM may have coincided with poorer clinical outcomes. However, given the small sample size, retrospective toxicity assessment and use of anthropometric rather than CT-derived estimates, no predictive conclusions can be drawn. These findings are hypothesis-generating and support further prospective evaluation of body composition alongside chemotherapy delivery. Retrospective CTCAE grading highlighted limitations in routine toxicity documentation. Standardised toxicity recording within electronic prescribing systems could improve consistency, support dose-modification decisions and facilitate future audit. [9]

Three service-improvement priorities emerge: routine monitoring of RDI as a quality indicator, standardised CTCAE documentation, and prospective evaluation of supportive care strategies to optimise chemotherapy delivery while maintaining treatment completion.

## Limitations

This study has several limitations. Its retrospective, single-centre design and small sample size (n=12) limited the analysis to descriptive findings. Toxicity grading was performed retrospectively, and lower-grade toxicities may therefore have been under-reported. Dose-capping decisions were not protocolised and may have varied according to individual clinical judgement. Finally, the relatively short study period and follow-up limited assessment of long-term oncological outcomes and precluded meaningful survival analysis.

Estimated LBM was derived from anthropometric equations rather than CT imaging, and observations relating to LBM should be regarded as exploratory and hypothesis-generating.

## Conclusion

This single-centre service evaluation found high completion of preoperative FLOT and progression to curative-intent surgery despite relative dose intensity below that reported in clinical trials. Although major pathological response was lower than in FLOT4-AIO, the contribution of reduced dose intensity cannot be determined from this small descriptive study. Prospective multicentre studies are needed to evaluate strategies that optimise chemotherapy delivery, treatment tolerability and relative dose intensity in routine NHS practice.

## Supporting information

Supplementary Appendix 1. ESMO-GROW flow diagram for real-world evidence study

## Data Availability

All data produced in the present study are available upon reasonable request to the authors. These will be shortly uploaded to zenodo (the data file)

## Ethics and Governance

This audit was conducted as a clinical governance service evaluation (CSCS/ONC/142) and did not require research ethics committee approval.

## Acknowledgements

We acknowledge Dr Kathryn Humphries and Dr Ahmed Mohamed for their insights and opinion on the final manuscript

## Funding

No specific funding was received for this work.

## Competing interests

None declared

## Data availability statement

Data are available from the corresponding author on reasonable request, subject to local governance approvals.

## Notes

### Competing Interest Statement

The authors have declared no competing interest.

### Author Declarations

No ethics approval was required, the service evaluation was registred with York and Scarborough clinical governance team and the dataset used for fully anonymised, no patient consent was required for this. The evaluation was registered with the hospital clinical governance department (CSCS/ONC/142).

## References

[1] Al-Batran SE, Homann N, Pauligk C, et al. Perioperative chemotherapy with fluorouracil plus leucovorin, oxaliplatin, and docetaxel versus fluorouracil or capecitabine plus cisplatin and epirubicin for locally advanced, resectable gastric or gastro-oesophageal junction adenocarcinoma (FLOT4): a randomised, phase 2/3 trial. Lancet 2019;393:1948–57. doi:10.1016/S0140-6736(18)32557-1.

[2] National Comprehensive Cancer Network. NCCN Clinical Practice Guidelines in Oncology: Gastric Cancer. Version 1.2026. Plymouth Meeting, PA: NCCN; 2026. Available from: https://www.nccn.org

[3] Geerts JFM, van der Zijden CJ, van der Sluis PC, et al. Perioperative chemotherapy for gastro-oesophageal or gastric cancer: anthracycline triplets versus FLOT. Cancers (Basel) 2024;16:1291. doi:10.3390/cancers16071291.

[4] Buonaiuto R, Martorana F, Marino P, et al. Association of neoadjuvant chemotherapy dose intensity with pathological complete response and event-free survival in HER2-negative early breast cancer. Eur J Cancer 2025;232:116102. doi:10.1016/j.ejca.2025.116102.

[5] Qi W, Wang X, Gan L, et al. The effect of reduced relative dose intensity of chemotherapy on outcomes in breast cancer patients. Sci Rep 2020;10:13241. doi:10.1038/s41598-020-70187-8.

[6] Wildiers H, Reiser M. Relative dose intensity of chemotherapy and its impact on outcomes in patients with early breast cancer or aggressive lymphoma. Crit Rev Oncol Hematol 2011;77:221–40. doi:10.1016/j.critrevonc.2010.02.002.

[7] Lavacchi D, Conticello C, Giommoni E, et al. Impact of relative dose intensity (RDI) of perioperative FLOT in resectable and locally advanced gastric and gastroesophageal junction adenocarcinoma. J Clin Oncol 2025;43(Suppl 16):e24089. . doi:10.1200/JCO.2025.43.16_suppl.e24089.

[8] Shayne M, Harvey RD, Lyman GH. Prophylaxis and treatment strategies for optimizing chemotherapy relative dose intensity. Expert Rev Anticancer Ther 2021;21:1145–59. doi:10.1080/14737140.2021.1941891.

[9] Sanyal A, Wellner D, Thomas J, et al. Standardization of toxicity-related dose modification in oncology electronic health record (EHR). J Clin Oncol 2022;40(Suppl 16):e18616. Abstract. doi:10.1200/JCO.2022.40.16_suppl.e18616.

[10] Janmahasatian S, Duffull SB, Ash S, et al. Quantification of lean bodyweight. Clin Pharmacokinet 2005;44:1051–65. doi:10.2165/00003088-200544100-00004.

[11] Griggs JJ, Bohlke K, Balaban EP, et al. Appropriate systemic therapy dosing for obese adult patients with cancer: ASCO guideline update. J Clin Oncol 2021;39:2037–48. doi:10.1200/JCO.21.00471.

[12] Williams G, Al-Obaidi M, Rower J, et al. Does oxaliplatin pharmacokinetics explain associations between body composition and chemotherapy toxicity risk in older adults with gastrointestinal cancers? J Clin Oncol 2021;39(Suppl 15):3095. Abstract. doi:10.1200/JCO.2021.39.15_suppl.3095.

[13] Graham MA, Lockwood GF, Greenslade D, et al. Clinical pharmacokinetics of oxaliplatin: a critical review. Clin Cancer Res 2000;6:1205–18. PMID:10778943.

[14] Prado CM, Baracos VE, McCargar LJ, et al. Body composition as an independent determinant of 5-fluorouracil-based chemotherapy toxicity. Clin Cancer Res 2007;13:3264– 68. doi:10.1158/1078-0432.CCR-06-3067.

[15] Drami I, Pring ET, Gould L, et al. Body composition and dose-limiting toxicity in colorectal cancer chemotherapy treatment: a systematic review of the literature. Could muscle mass be the new body surface area in chemotherapy dosing? Clin Oncol (R Coll Radiol) 2021;33:e540–52. doi:10.1016/j.clon.2021.05.011.

[16] Denduluri N, Patt DA, Wang Y, et al. Dose delays, dose reductions, and relative dose intensity in patients with cancer who received adjuvant or neoadjuvant chemotherapy in community oncology practices. J Natl Compr Canc Netw 2015;13:1383–93. doi:10.6004/jnccn.2015.0166.

[17] Puri T, Blake GM. Comparison of ten predictive equations for estimating lean body mass with dual-energy X-ray absorptiometry in older patients. Br J Radiol 2022;95:20210378. doi:10.1259/bjr.20210378.

[18] Castelo-Branco L, Booth CM, Del Paggio JC, et al. ESMO Guidance for Reporting Oncology real-World evidence (GROW). Ann Oncol 2023;34. doi:10.1016/j.annonc.2023.10.001.

[19] Al-Batran SE, Hofheinz RD, Pauligk C, et al. Histopathological regression after neoadjuvant docetaxel, oxaliplatin, fluorouracil and leucovorin versus epirubicin, cisplatin and fluorouracil or capecitabine in patients with resectable gastric or gastro-oesophageal junction adenocarcinoma (FLOT4-AIO): results from the phase 2 part of a multicentre, open-label, randomised phase 2/3 trial. Lancet Oncology 2016;17:1697–708. doi:10.1016/S1470-2045(16)30531-9.

