## Supplementary Appendix 1. ESMO-GROW flow diagram for real-world evidence study for "Real-world perioperative FLOT delivery in gastric and gastro-oesophageal junction adenocarcinoma: a single-centre service evaluation in a District General Hospital in the UK"

**DATA SOURCE**

Dataset 1
Electronic Patient Record
District General Hospital NHS Trust
Individual patients (n=13)

Dataset 2
Electronic Chemotherapy Prescribing System
Individual patients (n=13)

↓

**DATA SOURCES LINKAGE OR MERGING**

Encounter variable(s):
Hospital identifier
NHS number
Treatment episode

↓

**ELIGIBILITY**

Cases identified:
Consecutive patients receiving perioperative FLOT
January–December 2024

n=13

↓

**EXCLUSION CRITERIA**

Dataset 1:
Discontinued treatment after one cycle (n=1)

Dataset 2:
None

↓

**CASES INCLUDED**

Cases included

n=12

↓

**FINAL DATA CLEANING**

Exclusion on data cleaning:
Misclassification (n=0)
Missing core variables (n=0)
Loss to follow-up (n=0)
Missing survival data (not applicable)
Others (n=0)

↓

**ANALYSIS**

Cases for analysis

n=12

Outcomes:
Treatment completion
Relative dose intensity
Dose modifications
Toxicity
Progression to surgery
Pathological response
Estimated lean body mass

Figure legend: ESMO-GROW flow diagram describing data sources, linkage, eligibility, exclusions, final data cleaning and final analysis cohort for this retrospective observational service evaluation.
